# Investigating naming error patterns after non-invasive brain stimulation and language treatment in persons with aphasia

**DOI:** 10.64898/2026.06.08.26354856

**Authors:** Myra J. Sydnor, Micah Alan Johnson, Becky Lammers, Jamie L. Murter, Martin Lindquist, Rajani Sebastian

## Abstract

**Background:** Transcranial direct current stimulation (tDCS) paired with behavioral language therapy can improve naming in persons with aphasia (PWA), yet naming errors persist. Little is known about how naming error patterns change after non-invasive brain stimulation is combined with language treatment.

**Aims:** To examine whether right cerebellar tDCS plus computerized aphasia therapy changes the types of naming errors in people with chronic aphasia across timepoints, and to determine whether effects differ by cerebellar tDCS polarity (anode vs. cathode).

**Methods and Procedures:** In a randomized, double-blind, sham-controlled, within-subject crossover study, we retrospectively analyzed behavioral data from 24 individuals with post-stroke aphasia. Each participant completed two 15-session intervention periods (3–5 sessions/week) with active cerebellar tDCS + computerized aphasia therapy and sham + computerized aphasia therapy, separated by a two-month washout. General linear models (GLMs) assessed longitudinal changes in six error types (semantic, phonological real word, phonological nonword, no response, mixed, unrelated) on an untrained picture naming task (Philadelphia Naming Test; PNT) and a trained task (Naming 80; N80). Additional GLMs evaluated polarity effects with 2 (Group: anode vs. cathode) × 2 (Treatment) interactions, and treatment-order effects with 2 (Group: tDCS-first vs. sham-first) × 2 (Treatment) interactions.

**Outcomes and Results:** Active cerebellar tDCS did not significantly change error types for trained items (N80). For untrained items (PNT), active tDCS reduced several error types relative to sham, with the clearest and most durable reduction in phonological nonword errors; more moderate reductions occurred for phonological real word and unrelated errors. Mixed errors showed a marginally opposite pattern, tending to increase after tDCS and decrease after sham.

Polarity analyses indicated broadly similar effects across anodal and cathodal stimulation overall, but only the anode group showed a reliable treatment effect for phonological nonword errors on the PNT. Treatment-order analyses revealed no significant order effects.

**Conclusions:** Our results indicate a shift in naming error types, particularly after tDCS treatment for the untrained naming task (PNT). These findings may help guide the course of treatment approaches of those with aphasia and what error naming pattern types may show changes post stroke when combining non-invasive brain stimulation and computerized aphasia therapy.

**Clinical Trial Registration:** Cerebellar Transcranial Direct Current Stimulation and Aphasia Treatment [NCT02901574]

## Introduction

Following a stroke, anomia or word-finding difficulty is pervasive and often a challenge in the recovery process for persons with aphasia (PWA; Goodglass & Wingfield, 1997) in which there are difficulties in lexical access and word retrieval (Li et al., 2025). Speech-language therapy is the primary intervention for anomia, and interventions which focus on anomia are an integral component of aphasia therapy (Wisenburn & Mahoney, 2009)). There are many approaches for treatment of anomia, including treatments that target underlying linguistic processes necessary for successful word retrieval (e.g., semantic, phonological). There is substantial evidence documenting the benefits of anomia treatment for word-retrieval deficits in post-stroke aphasia including several reviews and meta-analysis that have shown beneficial effects of anomia treatment (e.g., Braun et al., 2025; Efstratiadou et al., 2018; Madden et al., 2017; Nickels, 2002; Sze et al., 2021; Wisenburn & Mahoney, 2009). A recent meta-analysis study focused on the efficacy of confrontational naming treatments for aphasia and found that syntactic cueing, errorless learning, and action observation methods have the highest effect size and are, therefore, the most effective. Additionally, the semantic feature analysis (SFA) and gestural methods are also effective interventions, considering the abundance of studies, high level of evidence, and substantial effect size (Yousefzade et al., 2025). However, individuals with chronic anomia may make slow progress to achieve relatively small gains, especially if treatment is not provided intensively (Barthel et al., 2008).

Evidence is growing that the add-on use of non-invasive brain stimulation techniques such as transcranial direct current stimulation (tDCS) along with behavioral aphasia therapy can aid in the recovery of naming in PWA (for reviews see Berube & Hillis, 2019; Biou et al., 2019; Breining & Sebastian, 2020; Elsner et al., 2020; Raymer & Johnson, 2024; Sebastian et al., 2020; Zettin et al., 2021). Studies, including our work, have shown that tDCS with anomia treatment, including computerized anomia treatment, can provide positive benefits in trained naming with generalization to untrained naming, discourse, and functional communication skills (Baker et al., 2010; Bonilha et al., 2024; Fridriksson et al., 2018; Holland & Crinion, 2012; Marangolo et al., 2014, 2018; Sebastian et al., 2020; Stockbridge et al., 2023). The main outcome measures in the majority of the tDCS investigations typically include naming accuracy on trained and untrained items. However, it is unclear how the patterns of naming errors change with tDCS, which has the potential to delineate the linguistic mechanisms underlying improvements following treatment.

Several behavioral aphasia treatment studies have explored changes in naming errors pre-treatment to post-treatment (e.g., Gordon, 2007; Hashimoto et al., 2013; Kendall et al., 2013; Kiran & Johnson, 2008; Kiran & Thompson, 2003; Li et al., 2025; Minkina et al., 2016, 2019). For example, Kendall and colleagues have done several studies to examine changes in naming errors of persons with chronic aphasia after phonomotor treatment (Kendall et al., 2013; Minkina et al., 2016, 2019). In the initial study, Kendall et al., (2013) examined improvement in lexical retrieval via confrontation naming error profiles of 10 PWA. In this study, participants improved in their ability to name trained items and maintained these improvements three months after treatment. Additionally, an analysis of word retrieval error type showed a trend toward a decrease in the number of omission errors on trained words and a trend toward an increase in the number of mixed errors on untrained words. In a larger study with 28 PWA (Minkina et al., 2019), participants showed improvement in whole-word naming accuracy on trained items and maintained their improvement at 3 months post-treatment. Treatment effects also generalized to untrained nouns at three months post-treatment. Additionally, incorrect responses were coded for error type, and error proportions of each error type (e.g., semantic, phonological, omission). Participants demonstrated a decrease in proportions of omission and description errors on trained items immediately post-treatment. The results of the error analysis suggest that a global (i.e., both lexical–semantic and phonological) change in lexical knowledge underlies the observed changes in confrontation naming accuracy following phonomotor treatment.

Other studies have focused on studying changes in error patterns following semantically based treatment for anomia. Hashimoto et al. (2013) used a cross-over design to compare a semantic treatment that emphasized word category membership with a treatment that emphasized semantic features. Six of their eight participants improved on trained items, with no obvious advantage for one treatment condition over another when analyzing response accuracy; however, a difference between groups was found when considering naming errors. Both treatments were associated with a decrease in omission errors, while only the categorical treatment was associated with an increase in semantic errors, demonstrating a potential difference in treatment-induced changes in the linguistic network that would have been missed with analyses focused only on the whole-word accuracy level. A recent study using a typicality-based semantic feature analysis (SFA) treatment in 30 individuals with chronic aphasia examined whether word production errors evolved 1) for the trained and untrained items vs. assessed items, and 2) for the typical untrained items vs. atypical untrained item (Li et al., 2025). Participants showed significant treatment and generalization effects, as evidenced by significant changes on error coding scores for both trained and untrained items relative to the assessed items. Word production errors for the trained items evolved from semantic/mixed errors to phonemic errors as expected given literature suggesting that targeting the semantic system will support improvements in the phonological system. Additionally, a significant change of word production errors was noted for the untrained stimuli, from semantic to mixed errors for the typical untrained stimuli and from mixed to phonemic errors for the atypical untrained stimuli, suggesting that typicality-based SFA treatment facilitates generalization to semantically related but untrained items.

Together, these results suggest an overall shift in the level of linguistic processing following treatment, encompassing changes in both lexical–semantic and phonological knowledge. However, it is not clear if the results would be similar for aphasia treatment combined with tDCS. The aim of this study was to examine changes in picture naming errors for trained and untrained items in PWA who underwent cerebellar tDCS and sham tDCS combined with computerized aphasia treatment. We hypothesized that there would be a significant difference in picture naming error types on trained and untrained naming post-treatment, 2-weeks post-treatment and 2-months post-treatment compared to pre-treatment for tDCS, but not for sham conditions. In addition, we hypothesized differential treatment effects (tDCS versus sham) depending on cerebellar tDCS polarity (anode versus cathode).

## Materials and Methods

### Participants

We retrospectively reviewed behavioral data for 24 participants (mean age= 64.4, four females) with chronic aphasia (mean time post onset = 46 months) due to left hemisphere stroke. Data were collected from a prior larger research study in which the behavioral and neural correlates of cerebellar tDCS in aphasia treatment were examined [Clinical Trial registration NCT02901574]. Most participants in the study had large left middle cerebral artery strokes. The demographic information of each participant is summarized in Table 1 and the participant recruitment flowchart is in Supplementary Figure S1. The inclusion criteria for participants were as follows: left hemisphere stroke; greater than 6 months post-stroke; at least 18 years or older; previously right-hand dominant; diagnosed with aphasia after evaluation using the short version of the Boston Diagnostic Aphasia Examination (BDAE; Goodglass et al., 2001); and able to obtain at least 65% accuracy on the aphasia treatment task screening version (details are in the Aphasia Treatment section; Kim et al., 2024; Sebastian et al., 2020). Exclusion criteria for participants included the following: lesion in the right cerebellum; history of brain surgery; seizures in the prior 12 months; sensitivity of the scalp as reported by the participant; and greater than 80% naming accuracy on the Philadelphia Naming Test (PNT; Kim et al., 2024; Roach et al., 1996; Sebastian et al., 2020). Written consent was provided by all the participants. Approval of the study was provided by the Institutional Review Board of the Johns Hopkins University School of Medicine, where all the data collection occurred.

**Table 1.**
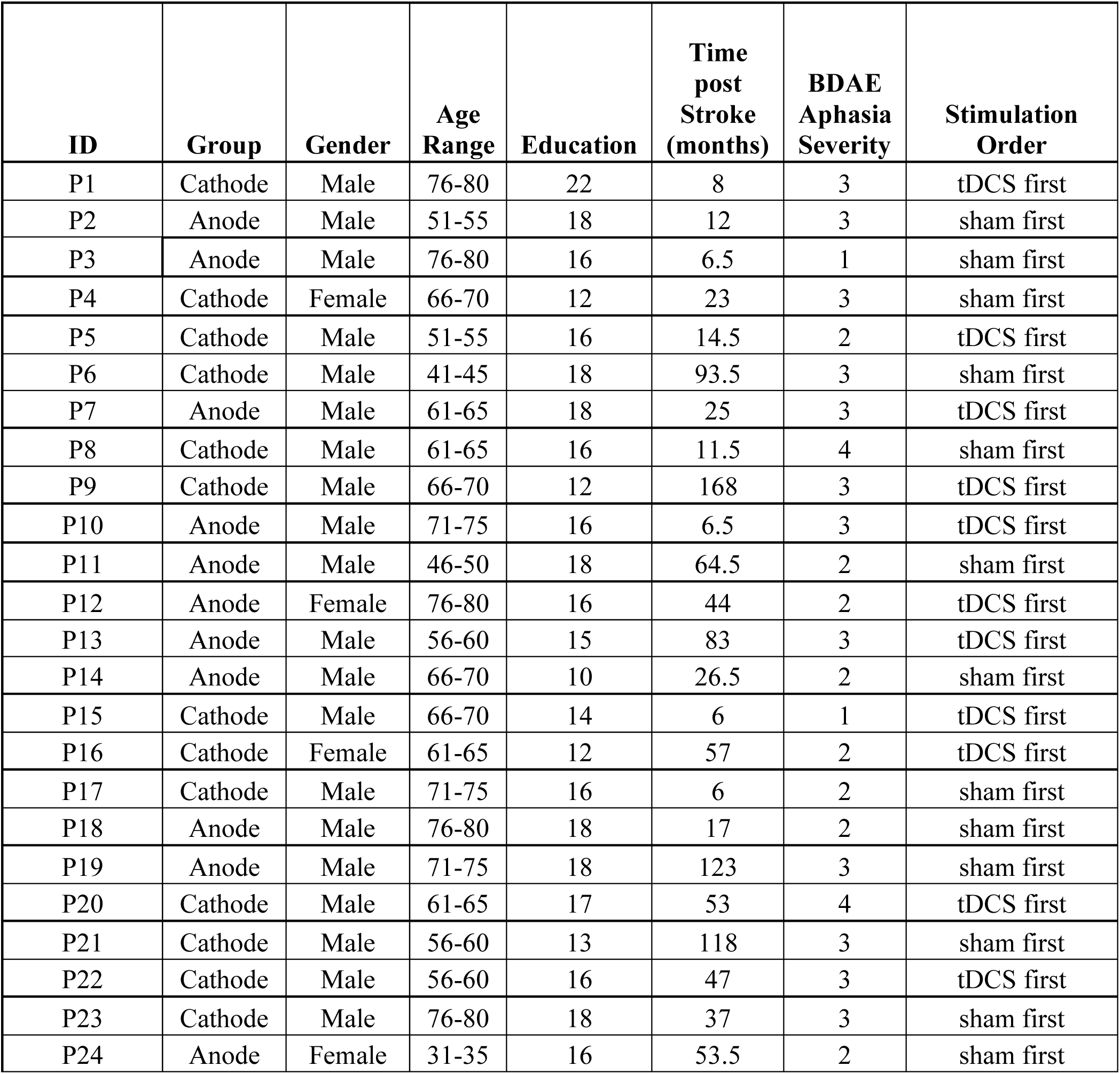
Demographics of participants; BDAE Aphasia Severity at baseline. BDAE: Boston Diagnostic Aphasia Examination.

### Study Design

This study used a randomized, double-blind, sham-controlled, within-subject crossover trial. There were two intervention periods of 15 (3-5 times/week) treatment sessions each, with cerebellar tDCS + computerized aphasia therapy and sham + computerized aphasia therapy, separated by a wash-out period of two months designed to minimize possible carryover effects from Phase 1 to Phase 2 (Kim et al., 2024; Sebastian et al., 2020). Each participant served as his or her own control. Participants who met eligibility criteria were randomly assigned using block randomization with a ratio of 1:1 to group anode or group cathode. Within each group, participants were randomly assigned to receive either ‘tDCS first then sham’ or ‘sham first then tDCS’. The order of whether the participant received ‘tDCS first’ ‘sham first’ was counterbalanced (Kim et al., 2024; Sebastian et al., 2020). Standardized language assessments were administered at each timepoint (pre-treatment, post-treatment, 2-weeks post-treatment, and 2-months post-treatment) for each phase. The 2-months post-treatment assessment for Phase 1 served as the Phase 2 pre-treatment assessment.

### Stimulation of cerebellar region

A Soterix 1×1 clinical trial device, with 2 mA of anodal or cathodal tDCS administered for 20 minutes to the right cerebellum and reference electrode placed on the right shoulder, was used (Sebastian et al., 2017, 2020; Turkeltaub et al., 2016). The electrodes were placed between two 5 cm x 5 cm sponges saturated with saline (Sebastian et al., 2020).

### Randomization and blinding

All of the participants as well as the research study team were blinded to the sequence in which each participant received tDCS stimulation or sham (Kim et al., 2024; Sebastian et al., 2020). A six-digit code was entered to account for blinding and for initiation of the stimulation. To achieve blinding for the participants, during the sham condition, stimulation was administered for the first 30 seconds in a ramp-up manner to foster the same sensation on the scalp as felt during the tDCS condition (Gandiga et al., 2006). Subsequently, the current was gradually reduced over 15 seconds.

### Computerized Aphasia Treatment

We used a computerized aphasia treatment focused on lexical-semantic processing which has demonstrated to improve naming in PWA (Baker et al., 2010; Fridriksson et al., 2018). Fridriksson and colleagues (Fridriksson et al., 2009, 2011, 2018) developed a computerized aphasia treatment program. This treatment program focused on 160 color pictures depicting low-medium and high frequency nouns that were randomly presented four times during the treatment with a semantic foil, phonological foil, unrelated word or the target word. Half of the pairs did represent a correct match (Sebastian et al., 2020). A laptop computer with two large response buttons was used. During treatment, a picture was shown on the laptop screen for about 2-5 seconds. Next, a video of the speaker’s face below the level of the nose was presented on the screen verbalizing a word that either matched or did not match the picture. The participant was then instructed to press the green response button if the word matched the picture or press the red response button if the word did not match the picture. Participants had 2-5 seconds to provide a response. Immediate feedback was given by either being shown a “smiley face” for correct responses or a “frowny face” for incorrect responses. The computer program did not advance to the next item until a response was recorded for the previous item.

To ensure comprehension of the treatment task, a pre-treatment screening, exactly matching the treatment task, but with 40 high-frequency words, was administered. Participants were provided with three opportunities to achieve 65% accuracy, a level of accuracy indicating that he or she understood the requirements of the task (Sebastian et al., 2020). The accuracy of the computerized aphasia treatment was recorded at the end of each session (Kim et al., 2024; Sebastian et al., 2020).

### Outcome Measures and Error Coding

Naming assessments were video recorded for transcribing, scoring, and error coding. Two outcome measures were used in this study: the number of correctly named pictures on the Naming 80 test (trained naming) and the number of correctly named pictures on the PNT (untrained naming). The Naming 80 was administered which focused on a subset of 80 pictures that were used during the treatment program. During the treatment program, there were 160 pictures which were used; however, only half of the treatment items were then selected in order to decrease assessment time at each timepoint. (Fridriksson et al., 2009; Sebastian et al., 2020).

Outcome measures were assessed before and after the end of the treatment, 2-weeks post-treatment and 2-months post-treatment for the tDCS and sham conditions. The 2-month follow-up for Phase 1 served as the baseline for the pre-treatment for Phase 2. Naming assessments were video recorded for transcribing, scoring, and error coding. First, items were scored as correct versus incorrect. Naming accuracy was scored based on general PNT scoring guidelines (Roach et al., 1996). After each word was coded for accuracy, any incorrectly named items were then further coded for error type using the following six categories: semantic (a real word that is similar in meaning to the target word, e.g., *book* for *library*), phonological real word (a real word similar in speech sound[s] to the target word, *mouse* for *mouth)*, phonological nonword (a nonword similar in speech sound[s] to the target word, e.g., *cordnet* for *carpet)*, mixed (a combination of sematic and phonological, e.g., *asparagus* for *apricot)*, no response (included no verbal response given or fragments, neologisms or nonwords [e.g., *deo* for stingray, “I don’t know”, or “no” provided], and unrelated responses (not related or similar to the target word, e.g., *vase* for *tuxedo)*.

### Reliability of Outcome Measures Scoring

To ensure reliability, 10% of the naming data were scored and coded by a second blind rater, and ratings were compared for agreement. A third rater was assigned to resolve any discrepancies that could not be resolved by the first and second raters, or to act as the second rater if the assigned second rater was also the tester. Naming accuracy, error coding and reliability were done by ASHA certified Speech-Language Pathologists. Discrepancies and ambiguous responses were also discussed at consensus meetings. Inter-rater reliability for naming accuracy (Cohen’s κ = 0.89) and type of response (Cohen’s κ = 0.85) were both strong (McHugh, 2012). The intraclass correlation coefficient (ICC) for the untrained naming (PNT) accuracy was 0.99 and for the trained naming (Naming 80) accuracy it was 0.98. The ICCs for the untrained naming (PNT) errors were as follows: semantic (0.99), phonological real word (0.94), phonological nonword (0.99), mixed (0.93), no response (0.99), and unrelated (0.93). The ICCs for the trained naming (Naming 80) errors were as follows: semantic (0.97), phonological real word (0.80), phonological nonword (0.99), mixed (0.86), no response (0.99), and unrelated (0.99).

## Statistical Methods

### Data Analysis

For all analyses, the dependent variable was the percentage change in naming error rate at each post-treatment timepoint relative to pre-treatment baseline (i.e., (post-pre)/pre), separately for each treatment phase (active tDCS or sham). If the baseline score was 0, then the difference score (post – pre) was used instead to avoid a division by zero error. We performed general linear models (GLM) to assess cerebellar tDCS treatment effects on longitudinal error changes separately for the six types of errors (semantic, phonological real word errors, phonological nonword errors, unrelated, mixed, and no response) for the trained (N80) and untrained (PNT) tasks. The primary independent variables included the within-subject categorical factors of Treatment (tDCS vs. sham) and Timepoint (post-treatment, 2-weeks post-treatment, 2-months post-treatment), modeled as either a 2 (Treatment) x 3 (Timepoint) interaction or as main effects (i.e., Treatment + Timepoint). The model also controlled for aphasia severity (i.e., BDAE overall aphasia severity) and the order of treatment phases (tDCS first then sham or sham first then tDCS).

We conducted two additional analyses. Additional GLM models tested cerebellar tDCS polarity effects with a 2 (Group: anode vs. cathode) x 2 (Treatment) interaction comparing treatment effects between anode and cathode groups, as well as between-subjects treatment phase order effects with a 2 (Group: tDCS first vs. sham first) x 2 (Treatment) interaction comparing treatment effects between treatment order groups. Both models controlled for aphasia severity and the model testing polarity effects also controlled for order effect. Because the limited sample size precluded testing the full Group (anode, cathode) x Treatment (tDCS, sham) x Timepoint (post-treatment, 2-wk post-treatment, 2-mo post-treatment) interaction within a GLM, nonparametric paired-samples t-tests (Wilcoxon signed-rank tests) of treatment effects (tDCS vs. sham) at each timepoint for different trained (N80) and untrained (PNT) error types were conducted within each group in order to qualitatively compare between groups.

All GLM analyses were performed in R Studio (Posit team, 2024) with glmmTMB (Brooks et al., 2017), emmeans or emtrends (Lenth, 2025) for post hoc tests (with FDR correction of p values at q = 0.05), eff_size (Lenth, 2025) for Cohen’s d effect sizes (Cohen, 1988) interpreted by standard convention (around 0.3 for small, 0.5 for medium, or 0.8 for large), DHARMa (Hartig et al., 2024) for model diagnostics, and ggplot2 (Wickham, 2016) for visualization. All nonparametric tests were performed in JASP (JASP Team, 2024; Love et al., 2019). The DVs were normalized with bestNormalize (Peterson, 2021; Peterson & Cavanaugh, 2020) to approximate a normal distribution and model diagnostics confirmed that a Gaussian distribution (identity link) and no random effects was appropriate in order to reduce or eliminate substantial problems with linearity, heteroscedasticity, homogeneity, dispersion, or outliers.

Multiple comparisons correction was performed with false discovery rate (FDR; q = 0.05; Benjamini & Hochberg, 1995) separately for all interaction or main effects across models and across post hoc tests with each model.

## Results

Tables 2 and 3 and Figures 1-4 show descriptive statistics of the naming error changes at each timepoint for tDCS and sham conditions for untrained and trained items.

**Figure 1.**
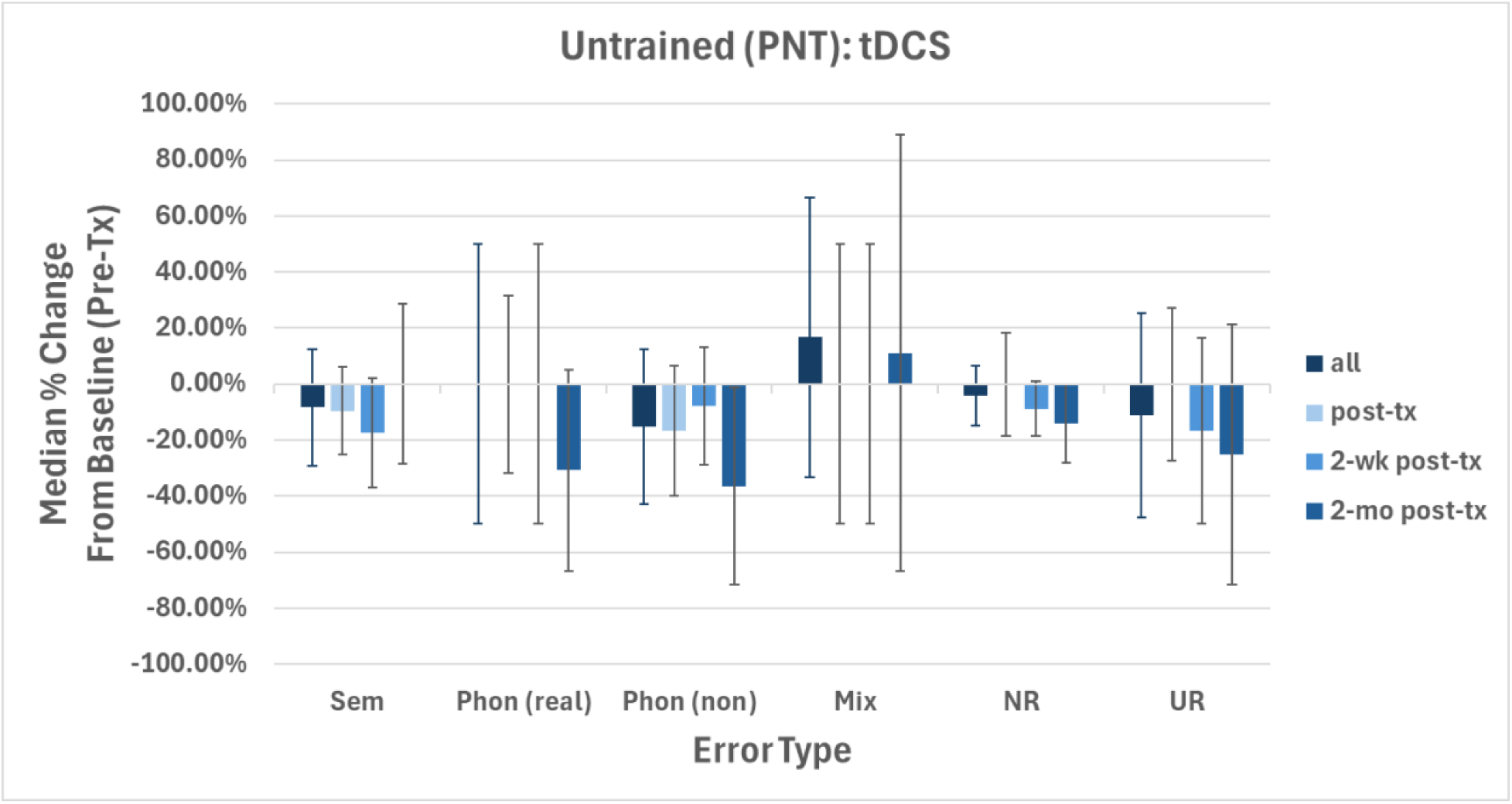
Descriptive statistics for the Untrained (PNT) task and the treatment condition (tDCS) showing naming error changes (medians and median absolute deviations) as percentages relative to baseline for each error type (“Sem” = semantic, “Phon (real)” = phonological real word, “Phon (non)” = phonological nonword, “Mix” = mixed, “NR” = no response, and “UR” = unrelated) and each post-treatment timepoint (“all” = total average, “post-tx” = immediate post-treatment, “2-wk post-tx” = 2 weeks post-treatment, “2-mo post-tx” = 2 months post-treatment).

**Figure 2.**
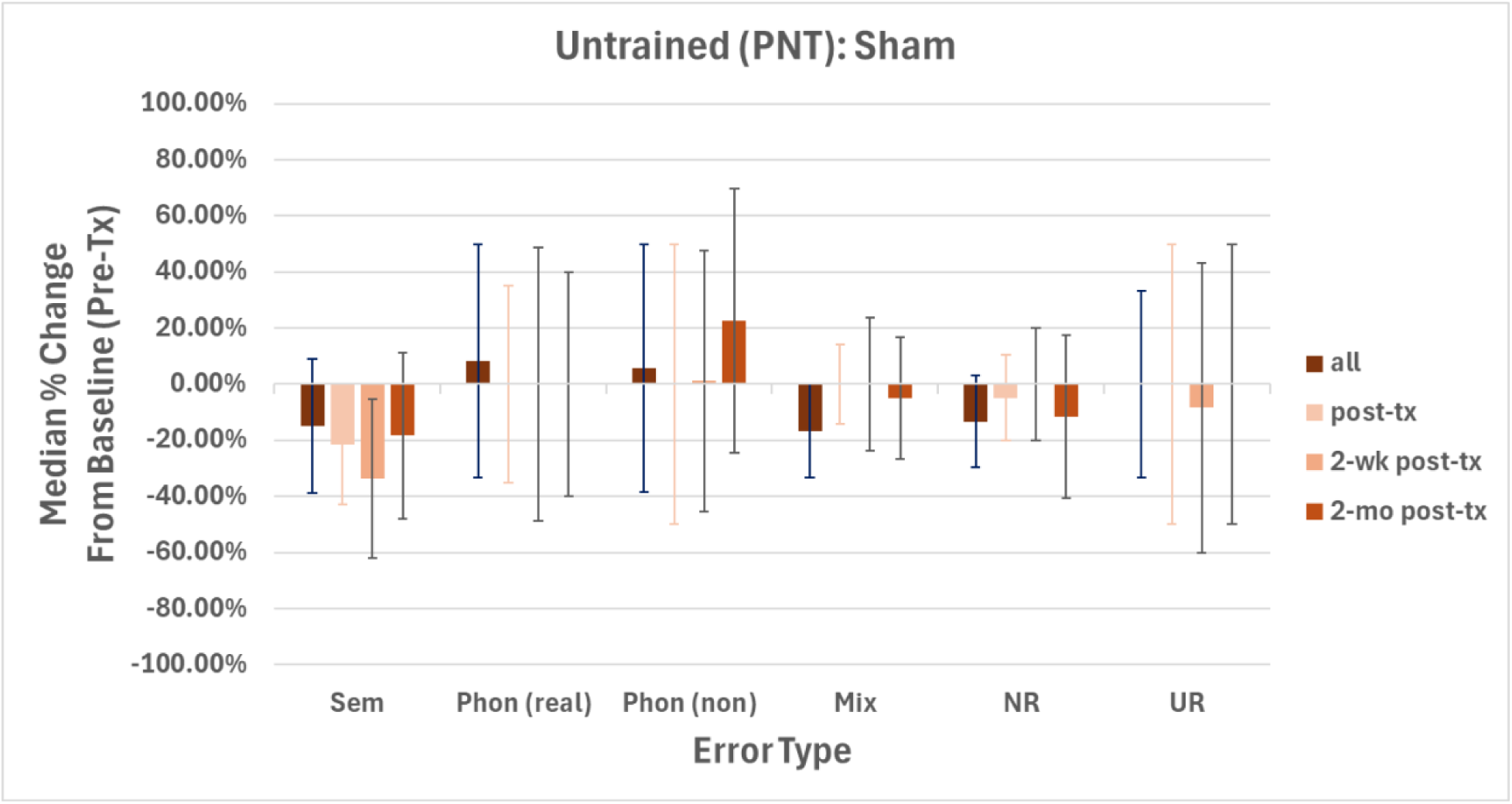
Descriptive statistics for the Untrained (PNT) task and the control condition (sham) showing naming error changes (medians and median absolute deviations) as percentages relative to baseline for each error type (“Sem” = semantic, “Phon (real)” = phonological real word, “Phon (non)” = phonological nonword, “Mix” = mixed, “NR” = no response, and “UR” = unrelated) and each post-treatment timepoint (“all” = total average, “post-tx” = immediate post-treatment, “2-wk post-tx” = 2 weeks post-treatment, “2-mo post-tx” =2 months post-treatment).

**Figure 3.**
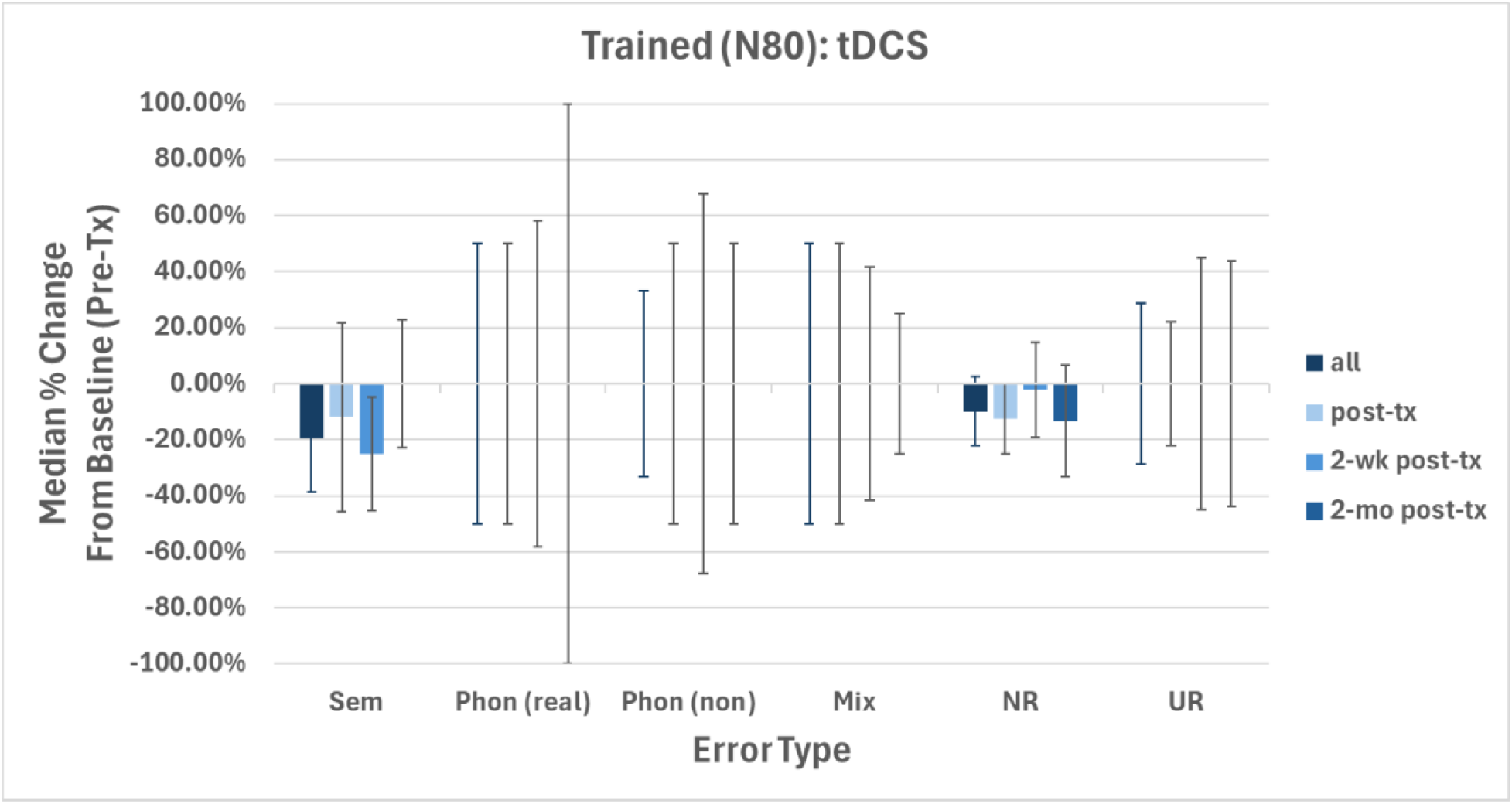
Descriptive statistics for the Trained (N80) task and the treatment condition (tDCS) showing naming error changes (medians and median absolute deviations) as percentages relative to baseline for each error type (“Sem” = semantic, “Phon (real)” = phonological real word, “Phon (non)” = phonological nonword, “Mix” = mixed, “NR” = no response, and “UR” = unrelated) and each post-treatment timepoint (“all” = total average, “post-tx” = immediate post-treatment, “2-wk post-tx” = 2 weeks post-treatment, “2-mo post-tx” = 2 months post-treatment).

**Figure 4.**
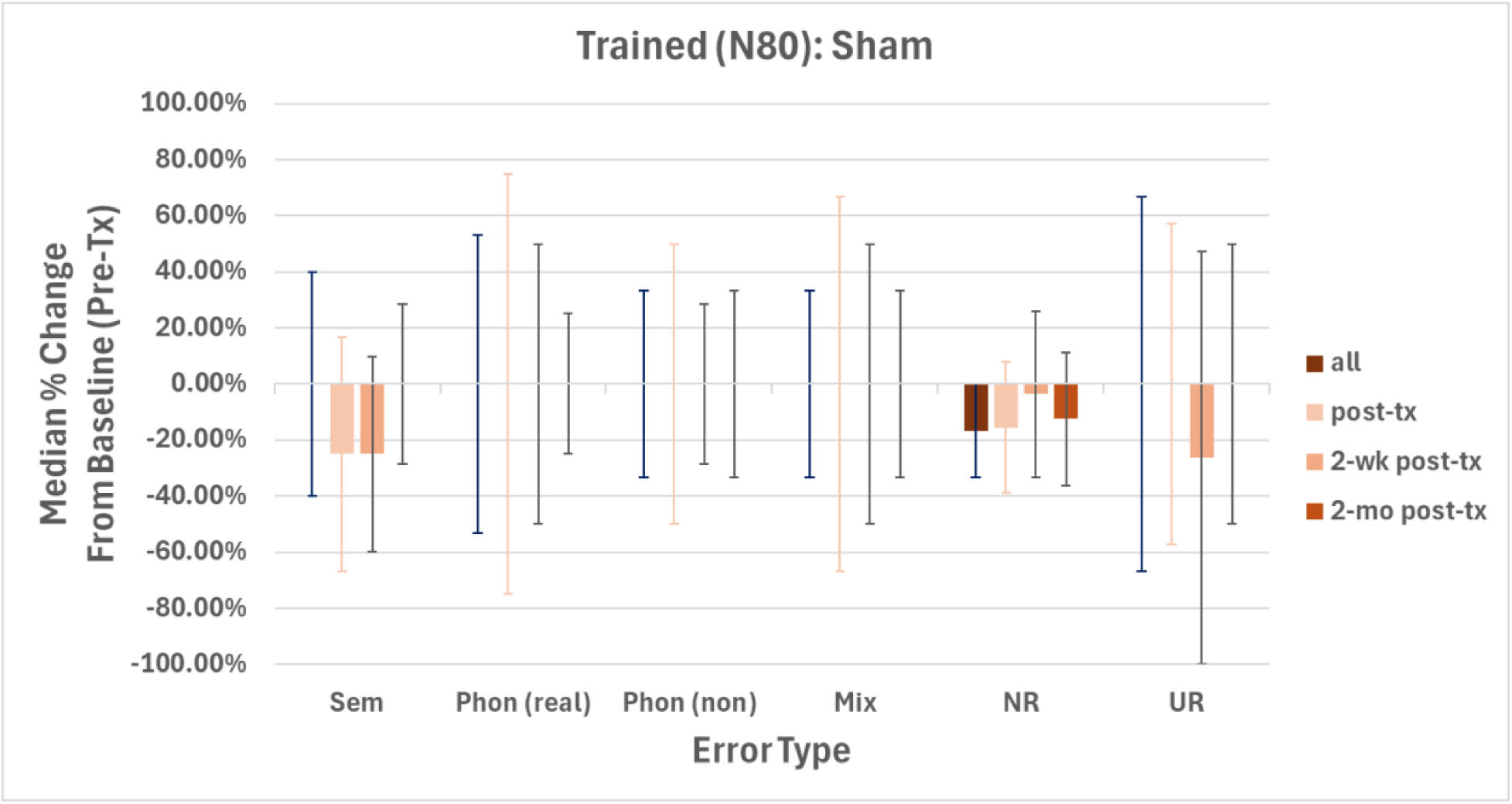
Descriptive statistics for the Trained (N80) task and the control condition (sham) showing naming error changes (medians and median absolute deviations) as percentages relative to baseline for each error type (“Sem” = semantic, “Phon (real)” = phonological real word, “Phon (non)” = phonological nonword, “Mix” = mixed, “NR” = no response, and “UR” = unrelated) and each post-treatment timepoint (“all” = total average, “post-tx” = immediate post-treatment, “2-wk post-tx” = 2 weeks post-treatment, “2-mo post-tx” = 2 months post-treatment).

**Table 2.**
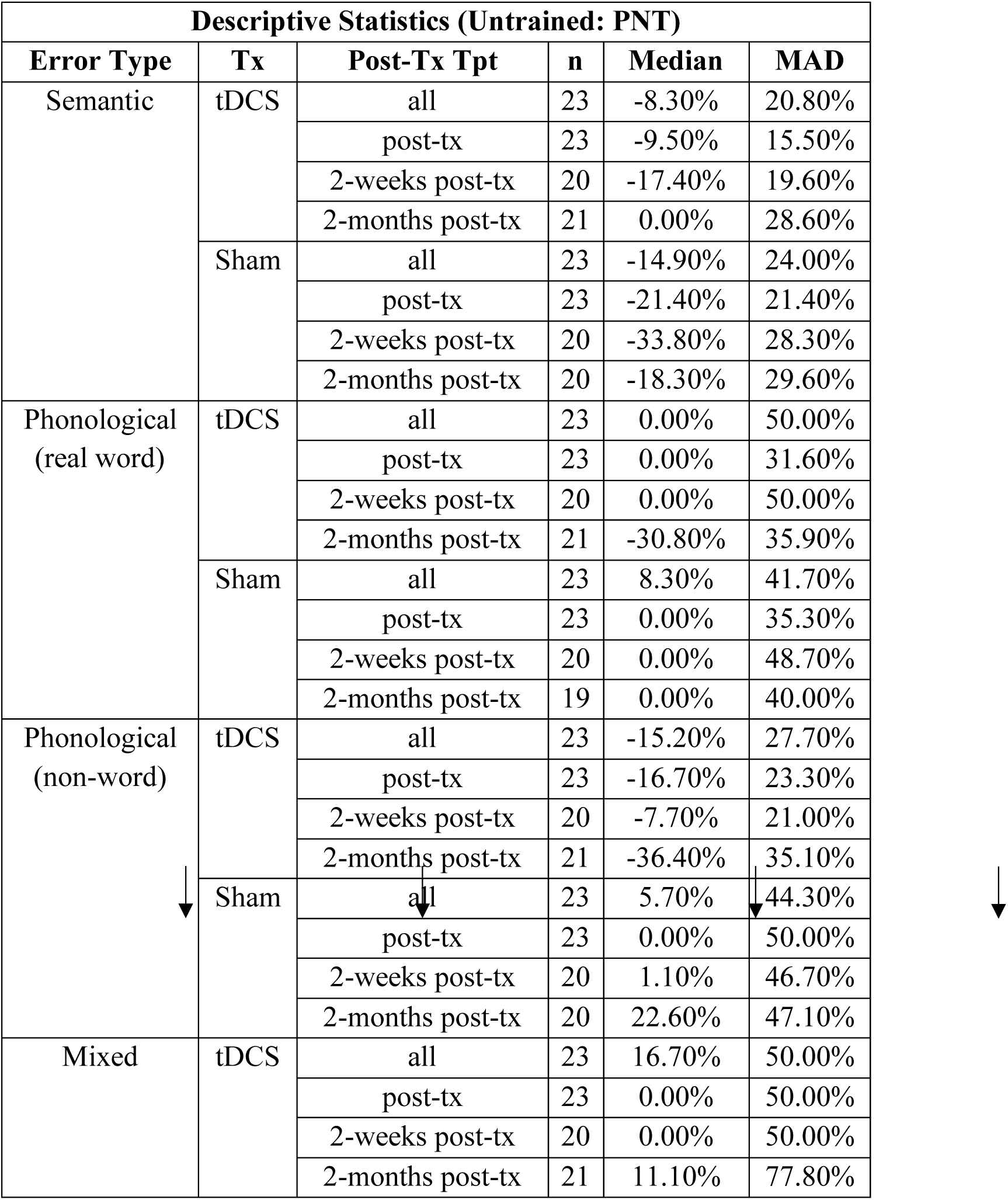
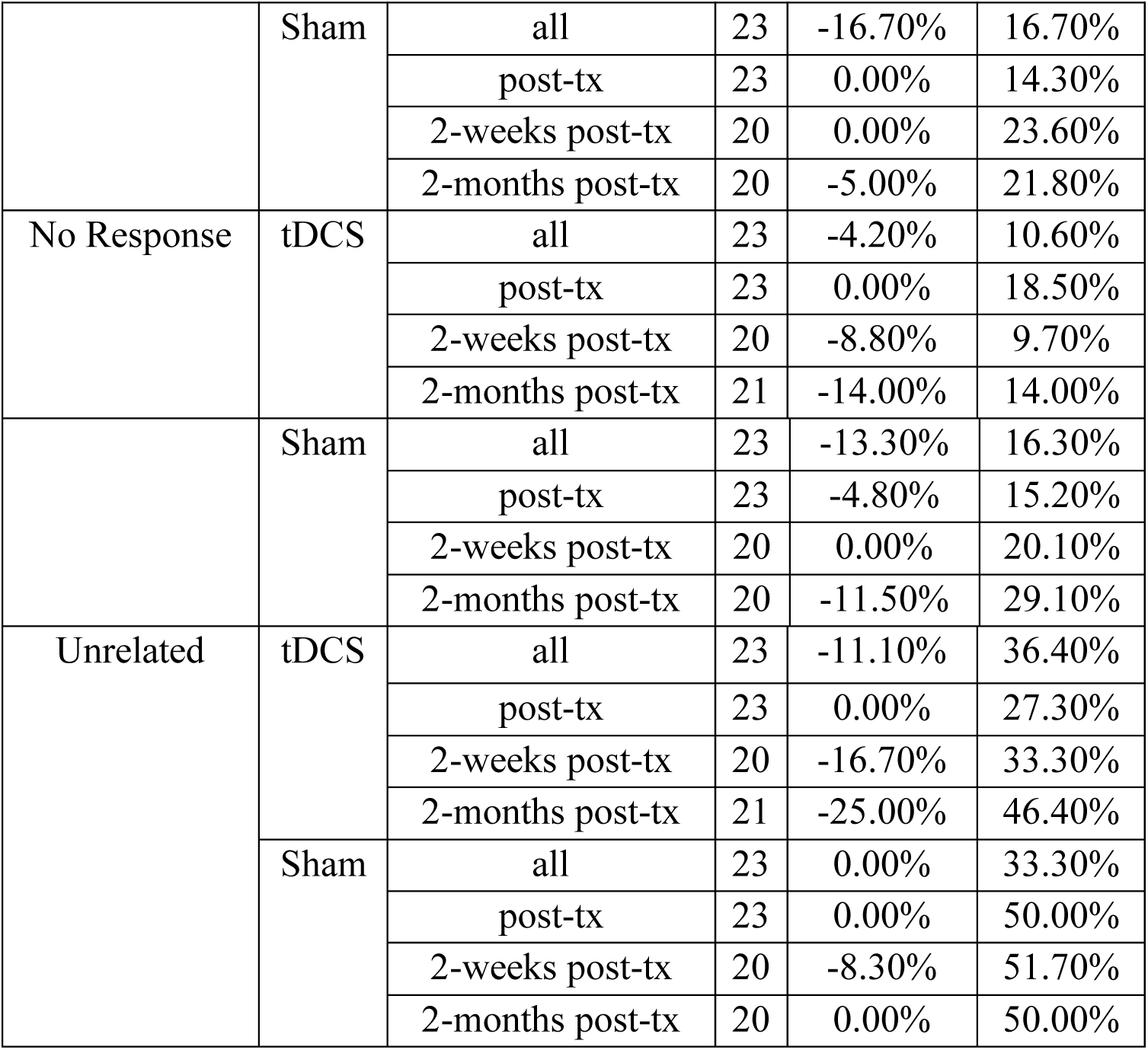
Descriptive statistics by treatment condition (active tDCS, sham) and post-treatment timepoints: all (overall average change,) post-treatment (post-tx), 2-weeks post-treatment (2-weeks post-tx), 2-months post-treatment(2-months post-tx) for each error type on the untrained naming task (Philadelphia Naming Test: PNT). MAD = Median Absolute Deviation. Values are percentage (%) change from the pre-treatment timepoint.

| **Descriptive Statistics (Untrained: PNT)** |  |  |  |  |  |
| --- | --- | --- | --- | --- | --- |
| **Error Type** | **Tx** | **Post-Tx Tpt** | **n** | **Median** | **MAD** |
| Semantic | tDCS | all | 23 | -8.30% | 20.80% |
|  |  | post-tx | 23 | -9.50% | 15.50% |
|  |  | 2-weeks post-tx | 20 | -17.40% | 19.60% |
|  |  | 2-months post-tx | 21 | 0.00% | 28.60% |
|  | Sham | all | 23 | -14.90% | 24.00% |
|  |  | post-tx | 23 | -21.40% | 21.40% |
|  |  | 2-weeks post-tx | 20 | -33.80% | 28.30% |
|  |  | 2-months post-tx | 20 | -18.30% | 29.60% |
| Phonological<br>(real word) | tDCS | all | 23 | 0.00% | 50.00% |
|  |  | post-tx | 23 | 0.00% | 31.60% |
|  |  | 2-weeks post-tx | 20 | 0.00% | 50.00% |
|  |  | 2-months post-tx | 21 | -30.80% | 35.90% |
|  | Sham | all | 23 | 8.30% | 41.70% |
|  |  | post-tx | 23 | 0.00% | 35.30% |
|  |  | 2-weeks post-tx | 20 | 0.00% | 48.70% |
|  |  | 2-months post-tx | 19 | 0.00% | 40.00% |
| Phonological<br>(non-word) | tDCS | all | 23 | -15.20% | 27.70% |
|  |  | post-tx | 23 | -16.70% | 23.30% |
|  |  | 2-weeks post-tx | 20 | -7.70% | 21.00% |
|  |  | 2-months post-tx | 21 | -36.40% | 35.10% |
|  | Sham | all | 23 | 5.70% | 44.30% |
|  |  | post-tx | 23 | 0.00% | 50.00% |
|  |  | 2-weeks post-tx | 20 | 1.10% | 46.70% |
|  |  | 2-months post-tx | 20 | 22.60% | 47.10% |
| Mixed | tDCS | all | 23 | 16.70% | 50.00% |
|  |  | post-tx | 23 | 0.00% | 50.00% |
|  |  | 2-weeks post-tx | 20 | 0.00% | 50.00% |
|  |  | 2-months post-tx | 21 | 11.10% | 77.80% |

| Descriptive Statistics (Untrained: PNT) |  |  |  |  |  |
| --- | --- | --- | --- | --- | --- |
| Error Type | Tx | Post-Tx Tpt | n | Median | MAD |
|  | Sham | all | 23 | -16.70% | 16.70% |
|  |  | post-tx | 23 | 0.00% | 14.30% |
|  |  | 2-weeks post-tx | 20 | 0.00% | 23.60% |
|  |  | 2-months post-tx | 20 | -5.00% | 21.80% |
| No Response | tDCS | all | 23 | -4.20% | 10.60% |
|  |  | post-tx | 23 | 0.00% | 18.50% |
|  |  | 2-weeks post-tx | 20 | -8.80% | 9.70% |
|  |  | 2-months post-tx | 21 | -14.00% | 14.00% |
|  | Sham | all | 23 | -13.30% | 16.30% |
|  |  | post-tx | 23 | -4.80% | 15.20% |
|  |  | 2-weeks post-tx | 20 | 0.00% | 20.10% |
|  |  | 2-months post-tx | 20 | -11.50% | 29.10% |
| Unrelated | tDCS | all | 23 | -11.10% | 36.40% |
|  |  | post-tx | 23 | 0.00% | 27.30% |
|  |  | 2-weeks post-tx | 20 | -16.70% | 33.30% |
|  |  | 2-months post-tx | 21 | -25.00% | 46.40% |
|  | Sham | all | 23 | 0.00% | 33.30% |
|  |  | post-tx | 23 | 0.00% | 50.00% |
|  |  | 2-weeks post-tx | 20 | -8.30% | 51.70% |
|  |  | 2-months post-tx | 20 | 0.00% | 50.00% |

**Table 3.** Descriptive statistics by treatment condition (active tDCS, sham) and post-treatment timepoints: all (overall average change), post-treatment (post-tx), 2-weeks post-treatment (2-weeks post-tx), 2-months post-treatment (2-months post-tx) for each error type on the trained naming task (N80). MAD = Median Absolute Deviation. Values are percentage (%) change from the pre-treatment timepoint.

| Descriptive Statistics (Trained: N80) |  |  |  |  |  |
| --- | --- | --- | --- | --- | --- |
| Error Type | Tx | Post-Tx Tpt | n | Median | MAD |
| Semantic | tDCS | all | 23 | -19.40% | 19.40% |
|  |  | post-tx | 23 | -11.80% | 33.70% |
|  |  | 2-weeks post-tx | 20 | -25.00% | 20.20% |
|  |  | 2-months post-tx | 21 | 0.00% | 22.70% |
|  | Sham | Average | 23 | 0.00% | 40.00% |
|  |  | post-tx | 23 | -25.00% | 41.70% |
|  |  | 2-weeks post-tx | 21 | -25.00% | 34.70% |
|  |  | 2-months post-tx | 21 | 0.00% | 28.60% |
| Phonological<br>(real word) | tDCS | all | 23 | 0.00% | 50.00% |
|  |  | post-tx | 23 | 0.00% | 50.00% |
|  |  | 2-weeks post-tx | 20 | 0.00% | 58.30% |
|  |  | 2-months post-tx | 21 | 0.00% | 100.00% |
|  | Sham | all | 23 | 0.00% | 53.30% |
|  |  | post-tx | 23 | 0.00% | 75.00% |
|  |  | 2-weeks post-tx | 21 | 0.00% | 50.00% |
|  |  | 2-months post-tx | 21 | 0.00% | 25.00% |
| Phonological<br>(non-word) | tDCS | all | 23 | 0.00% | 33.30% |
|  |  | post-tx | 23 | 0.00% | 50.00% |
|  |  | 2-weeks post-tx | 20 | 0.00% | 67.90% |
|  |  | 2-months post-tx | 21 | 0.00% | 50.00% |
|  | Sham | all | 23 | 0.00% | 33.30% |
|  |  | post-tx | 23 | 0.00% | 50.00% |
|  |  | 2-weeks post-tx | 21 | 0.00% | 28.60% |
|  |  | 2-months post-tx | 21 | 0.00% | 33.30% |
| Mixed | tDCS | all | 23 | 0.00% | 50.00% |
|  |  | post-tx | 23 | 0.00% | 50.00% |
|  |  | 2-weeks post-tx | 20 | 0.00% | 41.70% |
|  |  | 2-months post-tx | 21 | 0.00% | 25.00% |
| Error Type | Tx | Post-Tx Tpt | n | Median | MAD |
| Mixed | Sham | all | 23 | 0.00% | 33.30% |
|  |  | post-tx | 23 | 0.00% | 66.70% |
|  |  | 2-weeks post-tx | 21 | 0.00% | 50.00% |
|  |  | 2-months post-tx | 21 | 0.00% | 33.30% |
| No Response | tDCS | all | 23 | -9.80% | 12.40% |
|  |  | post-tx | 23 | -12.50% | 12.50% |
|  |  | 2-weeks post-tx | 20 | -2.20% | 16.90% |
|  |  | 2-months post-tx | 21 | -13.30% | 20.00% |
|  | Sham | all | 23 | -16.70% | 16.70% |
|  |  | post-tx | 23 | -15.60% | 23.30% |
|  |  | 2-weeks post-tx | 21 | -3.70% | 29.60% |
|  |  | 2-months post-tx | 21 | -12.50% | 23.80% |
| Unrelated | tDCS | all | 23 | 0.00% | 28.60% |
|  |  | post-tx | 23 | 0.00% | 22.20% |
|  |  | 2-weeks post-tx | 20 | 0.00% | 45.00% |
|  |  | 2-months post-tx | 21 | 0.00% | 43.80% |
|  | Sham | all | 23 | 0.00% | 66.70% |
|  |  | post-tx | 23 | 0.00% | 57.10% |
|  |  | 2-weeks post-tx | 21 | -26.30% | 73.70% |
|  |  | 2-months post-tx | 21 | 0.00% | 50.00% |

### Treatment Effects (tDCS Polarity Groups Combined)

#### Trained task (N80)

The primary research hypothesis was that active cerebellar tDCS treatment, relative to sham, would reduce the overall number and type of naming errors over time. For the trained (N80) task, however, there were no significant treatment effects as main effects or interacting with timepoint for any of the error types (see Table 4): semantic (Treatment x Timepoint: F(2, 120) = 0.13, p.fdr = 0.9725; Treatment: F(1, 122) = 0.03, p.fdr = 0.9894), phonological real word (Treatment x Timepoint: F(2, 120) = 0.03, p.fdr = 0.9725; Treatment: F(1, 122) = 0.00, p.fdr = 0.9894), phonological nonword (Treatment x Timepoint: F(2, 120) = 0.21, p.fdr = 0.9725; Treatment: F(1, 122) = 0.19, p.fdr = 0.9894), mixed (Treatment x Timepoint: F(2, 120) = 0.36, p.fdr = 0.9725; Treatment: F(1, 122) = 0.93, p.fdr = 0.9894), no response (Treatment x Timepoint: F(2, 120) = 0.52, p.fdr = 0.9725; Treatment: F(1, 122) = 0.34, p.fdr = 0.9894), and unrelated (Treatment x Timepoint: F(2, 120) = 1.29, p.fdr = 0.9725; Treatment: F(1, 122) = 2.65, p.fdr = 0.6389). The lack of treatment effects was due to the lack of substantial changes relative to baseline or relative to sham (for all post hoc test results, see Supplemental Tables S1a-S1b).

**Table 4.** Trained (N80) task. GLM results of the main effect of treatment (Tx) and interaction between treatment and timepoints (Tx x Tpt) for each error type (“Sem” = semantic, “Phon (real)” = phonological real word, “Phon (non)” = phonological nonword, “Mix” = mixed, “NR” = no response, and “UR” = unrelated).

| Task | Error type | Interaction (Tx x Tpt) |  |  |  |  | Main effect (Tx) |  |  |  |  |
| --- | --- | --- | --- | --- | --- | --- | --- | --- | --- | --- | --- |
|  |  | df1 | df2 | F | p | p.fdr | df1 | df2 | F | p | p.fdr |
| N80 | Sem | 2 | 120 | 0.13 | 0.8763 | 0.9725 | 1 | 122 | 0.03 | 0.8670 | 0.9894 |
|  | Phon (real) | 2 | 120 | 0.03 | 0.9725 | 0.9725 | 1 | 122 | 0.00 | 0.9894 | 0.9894 |
|  | Phon (non) | 2 | 120 | 0.21 | 0.8106 | 0.9725 | 1 | 122 | 0.19 | 0.6634 | 0.9894 |
|  | Mix | 2 | 120 | 0.36 | 0.6961 | 0.9725 | 1 | 122 | 0.93 | 0.3379 | 0.9894 |
|  | NR | 2 | 120 | 0.52 | 0.5943 | 0.9725 | 1 | 122 | 0.34 | 0.5613 | 0.9894 |
|  | UR | 2 | 120 | 1.29 | 0.2798 | 0.9725 | 1 | 122 | 2.65 | 0.1065 | 0.6389 |

#### Untrained task (PNT)

Some of the untrained (PNT) task error types showed evidence of treatment effects (see Table 5 for Treatment main effect and Treatment x Timepoint interactions and see Supplemental Tables S2a-S2b for full results of all post hoc tests).

**Table 5.** Untrained (PNT) task. GLM results of the main effect of treatment (Tx) and interaction between treatment and timepoints (Tx x Tpt) for each error type (“Sem” = semantic, “Phon (real)” = phonological real word, “Phon (non)” = phonological nonword, “Mix” = mixed, “NR” = no response, and “UR” = unrelated).

| Task | DV | Interaction (Tx * Tpt) |  |  |  |  | Main effect (Tx) |  |  |  |  |
| --- | --- | --- | --- | --- | --- | --- | --- | --- | --- | --- | --- |
|  |  | df1 | df2 | F | p | p.fdr | df1 | df2 | F | p | p.fdr |
| PNT | Sem | 2 | 118 | 0.36 | 0.7021 | 0.7284 | 1 | 120 | 1.61 | 0.2065 | 0.2478 |
|  | Phon (real) | 2 | 117 | 0.53 | 0.5926 | 0.7284 | 1 | 119 | 5.59 | 0.0197 | 0.0395 |
|  | Phon (non) | 2 | 118 | 2.27 | 0.1077 | 0.6463 | 1 | 120 | 15.60 | 0.0001 | 0.0008 |
|  | Mix | 2 | 118 | 0.62 | 0.5386 | 0.7284 | 1 | 120 | 3.88 | 0.0510 | 0.0766 |
|  | NR | 2 | 118 | 0.74 | 0.4790 | 0.7284 | 1 | 120 | 0.07 | 0.7897 | 0.7897 |
|  | UR | 2 | 118 | 0.32 | 0.7284 | 0.7284 | 1 | 120 | 5.71 | 0.0185 | 0.0395 |

For PNT phonological nonword errors, there was a significant main effect of Treatment (F(1, 120) = 15.60, p.fdr = 0.0008) with a moderately large effect size (d =-0.70, 95% CI [-1.05,-0.35]), averaging across all post-treatment timepoints, such that errors significantly decreased after tDCS (t(120) =-2.74, p.fdr = 0.0070) but significantly increased after sham (t(120) = 2.82, p.fdr = 0.0070). Although the Treatment x Timepoint interaction was not significant (F(2, 118) = 2.27, p.fdr = 0.6463), planned post hoc comparisons indicated that the main treatment effect was driven by the two months post-treatment timepoint, which showed a significant decrease in errors after tDCS (t(118) =-3.05, p.fdr = 0.0085) in contrast to a significant increase after sham (t(118) = 2.65, p.fdr = 0.0277), resulting in a significant treatment effect (tDCS vs. sham: t(118) =-4.03, p.fdr = 0.0003) with a large and reliable effect size (d =-1.26, 95% CI [-1.90,-0.62]). The earlier timepoints (immediate and 2 weeks post-treatment) showed numerical differences consistent with this pattern but no significant effects (see Supplemental Tables S2a-S2b).

For PNT phonological real word errors, the Treatment x Timepoint interaction was not significant (F(2, 117) = 0.53, p.fdr = 0.7284), but there was a significant main effect of Treatment (F(1, 119) = 5.86, p.fdr = 0.0395) with a moderate effect size (d =-0.42, 95% CI [-0.78,-0.07]), averaging across all post-treatment timepoints, such that errors tended to decrease after tDCS (t(119) =-1.63, p.fdr = 0.1053) but tended to increase after sham (t(119) = 1.69, p.fdr = 0.1053). For PNT unrelated errors, the Treatment x Timepoint interaction was not significant (F(2, 118) = 0.32, p.fdr = 0.7284), but there was a significant main effect of Treatment (F(1, 120) = 5.71, p.fdr = 0.0395) with a moderate effect size (d =-0.42, 95% CI [-0.78,-0.07]), averaging across all post-treatment timepoints, such that errors tended to decrease after tDCS (t(120) = - 1.84, p.fdr = 0.1290) but tended to increase after sham (t(120) = 1.53, p.fdr = 0.1290). For PNT mixed errors, the Treatment x Timepoint interaction was not significant (F(2, 118) = 0.62, p.fdr = 0.7284), but there was a marginally significant main effect of Treatment (F(1, 120) = 3.88, p.fdr = 0.0766) with a moderate effect size (d = 0.35, 95% CI [-0.001, 0.70]), averaging across all post-treatment timepoints, such that errors tended to increase after tDCS (t(120) = 1.44, p.fdr = 0.1817) but tended to decrease after sham (t(120) =-1.34, p.fdr = 0.1817). For PNT semantic errors, neither the main effect of Treatment (F(2, 120) = 1.61, p.fdr = 0.2478) or the Treatment x Timepoint interaction (F(2, 118) = 0.36, p.fdr = 0.7284) was significant.

### Comparing Treatment Effects between tDCS Polarity Groups

#### Trained task (N80)

For the trained (N80) task, none of the error types showed any significant differential treatment effects between anodal and cathodal polarity groups (i.e., tDCS Polarity Group x Treatment interaction) when averaging across timepoints: semantic (F(1, 122) = 0.00, p.fdr = 0.9748), phonological real word (F(1, 122) = 0.71, p.fdr = 0.7334), phonological nonword (F(1, 122) = 0.33, p.fdr = 0.7334), mixed (F(1, 122) = 1.99, p.fdr = 0.4825), no response (F(1, 122) = 0.26, p.fdr = 0.7334), unrelated (F(1, 122) = 4.91, p.fdr = 0.1713). There were also no significant differential treatment effects between polarity groups within separate timepoints based on nonparametric paired t-tests (see Supplemental Tables S3a-S3b).

#### Untrained task (PNT)

There was a significant Polarity Group (anode, cathode) x Treatment (tDCS, sham) interaction, averaging across all post-treatment timepoints, for the untrained phonological nonword errors (F (1, 120) = 9.46, p.fdr = 0.0156). Within the anode group, errors significantly decreased after tDCS (t(120) =-4.06, p.fdr = 0.0002) but significantly increased after sham (t(120) = 2.97, p.fdr = 0.0036), resulting in a significant treatment effect (tDCS vs. sham: t(120) =-5.06, p.fdr < 0.0001) with a large effect size (d =-1.32, 95% CI [-1.86,-0.78]). Planned comparisons of treatment effects (tDCS vs. sham) by timepoint, using nonparametric paired t-tests, revealed that the overall treatment effect in the anodal group was driven primarily by a significant error reduction at the 2-months post-treatment timepoint (W = 0.00, Z =-2.67, p.fdr = 0.0120) and also marginally significant reductions at the 2-weeks post-treatment (W = 5.00, Z =-1.82, p.fdr = 0.0800) and immediate post-treatment (W = 6.00, Z = - 1.96, p.fdr = 0.0800) timepoints. Within the cathode group, there was a nonsignificant decrease after tDCS (t(120) =-0.14, p.fdr = 0.8856) and a nonsignificant increase after sham (t(120) = 1.13, p.fdr = 0.5174), resulting in a nonsignificant treatment effect (tDCS vs. sham: t(120) = - 0.92, p.fdr = 0.3594) with a small and unreliable effect size (d =-0.22, 95% CI [-0.71, 0.26]). In other words, only the anode group showed a reliable treatment effect for untrained phonological nonword errors, driven by primarily the 2-months post-treatment timepoint.

None of the error types showed any significant differential treatment effects between anodal and cathodal polarity groups (i.e., tDCS Polarity Group x Treatment interaction) when averaging across timepoints: semantic (F(1, 120) = 1.75, p.fdr = 0.5672), phonological real word (F(1, 119) = 0.09, p.fdr = 0.8246), mixed (F(1, 120) = 0.20, p.fdr = 0.8246), no response (F(1, 120) = 0.20, p.fdr = 0.8246), unrelated (F(1, 120) = 0.05, p.fdr = 0.8246). There were also no other significant differential treatment effects between polarity groups within separate timepoints based on nonparametric paired t-tests (see Supplemental Tables S4a-S4b).

### Comparing Treatment Effects between Treatment Order Groups

#### Trained task (N80)

For the trained (N80) task, none of the error types showed any significant differential treatment effects between treatment order groups (tDCS-first vs. sham-first) (i.e., Order Group x Treatment interaction) when averaging across timepoints: semantic (F(1, 123) = 2.70, p.fdr = 0.1547), phonological real word (F(1, 123) = 4.40, p.fdr = 0.1139), phonological nonword (F(1, 123) = 3.69, p.fdr = 0.1139), mixed (F(1, 123) = 1.44, p.fdr = 0.2797), no response (F(1, 123) = 4.53, p.fdr = 0.1139), unrelated (F(1, 123) = 0.36, p.fdr = 0.5473).

#### Untrained task (PNT)

Similarly, for the untrained (PNT) task, none of the error types showed any significant differential treatment effects between treatment order groups (tDCS-first vs. sham-first) (i.e., Order Group x Treatment interaction) when averaging across timepoints: semantic (F(1, 121) = 1.10, p.fdr = 0.4463), phonological real word (F(1, 120) = 2.03, p.fdr = 0.4463), phonological nonword (F(1, 121) = 3.78, p.fdr = 0.3251), mixed (F(1, 121) = 1.19, p.fdr = 0.4463), no response (F(1, 121) = 0.00, p.fdr = 0.9853), unrelated (F(1, 121) = 0.00, p.fdr = 0.9853).

## Discussion

The current study investigated changes in picture naming errors for trained (N80) and untrained (PNT) items in PWA who received cerebellar tDCS and sham tDCS combined with computerized aphasia treatment. The primary research hypothesis was that active cerebellar tDCS treatment, relative to sham, would reduce the overall number and type of naming errors across various post-treatment timepoints. This hypothesis was partially supported. We found no significant effects of cerebellar tDCS on error types for trained items. In contrast, for untrained items, cerebellar tDCS reduced several error types relative to sham, with the clearest and most durable effect for phonological nonword errors. More moderate treatment effects were also observed for phonological real-word errors and unrelated errors. Mixed errors showed a marginal pattern in the opposite direction, tending to increase after tDCS while decreasing after sham. We also hypothesized differential treatment effects (tDCS vs. sham) depending on cerebellar tDCS polarity (anode vs. cathode). This hypothesis was also partially supported by polarity specific effect for the untrained task but not for the trained task. For the untrained task (PNT), only the anode group showed a reliable treatment effect for phonological nonword errors, the analysis that examined order effects, i.e., whether there was any difference in the treatment due to the order showed no significant effects.

The results of this analysis are in line with previous behavioral aphasia treatment studies that have explored changes in naming errors pre-treatment to post-treatment (Kendall et al., 2013; Li et al., 2025; Minkina et al., 2016, 2019).The results of error analysis are also in line with the results of naming accuracy data of Sebastian et al., 2020, where we found no significant effect of cerebellar stimulation for trained naming (N80), but a significant effect of cerebellar stimulation for untrained naming (PNT). Although cerebellar tDCS did not significantly change the distribution of picture naming error types for trained items, error patterns shifted from pre-to post-treatment and at follow-up time points. Across time points and in both the tDCS and sham conditions, omissions (no response) and semantic errors were the most frequent errors on trained items. Among error types, semantic errors showed the largest change with treatment, both with computerized aphasia treatment plus tDCS and with treatment alone. The predominance of pre-treatment semantic and omission errors suggests difficulty accessing the lemma in this group of PWA (Dell et al., 1997). In interactive activation accounts, semantically related words share features, so activation spreads from the target to its neighbors; in the presence of noise, a semantic neighbor may be selected, yielding a semantic error (Dell et al., 1997). Omissions likely occur when candidates at the semantic level fail to reach the selection threshold (Dell et al., 2004).

An explanation for the lack of cerebellar tDCS effects on trained naming errors data, consistent with Sebastian et al., 2020, is that the role the posterolateral cerebellum plays in language processing depends on task demands (Boehringer et al., 2013; Marangolo et al., 2018; Pope & Miall, 2012, 2014; Stoodley et al., 2010, 2012). Thus, it is likely that the right cerebellum contributes most when language behaviors are being actively acquired and optimized, and much less once stimulus–response mappings are consolidated, and execution becomes relatively automatic. In our study, trained items were repeatedly practiced with explicit feedback in a picture–word matching task, likely strengthening item-specific associations by post-treatment. Thus, lack of significant effects for trained naming could be due to the treatment task becoming more automatic and easier to perform.

The order that PWA received cerebellar tDCS versus sham likely contributed as well. In Sebastian et al., 2020, naming accuracy analysis, trained-item benefits were most evident when tDCS was delivered in the first phase, consistent with a larger cerebellar role early in learning. Even though the present error analysis did not show significant order effects, the broader pattern is consistent with cerebellar tDCS effects waning as tasks become more efficient. Our analysis, i.e., using the “change in naming error rate” approach reduced the carryover effect across phases, but it cannot remove consolidation of strategies and general task familiarity acquired in phase 1. Such consolidation, and occasional ceiling performance at the start of phase 2 for trained items, can compress between-condition differences and further obscure tDCS effects on trained naming errors.

For untrained naming (PNT), cerebellar tDCS produced a shift in several error types relative to sham. Like trained items, in both the tDCS and sham conditions, omissions (no response) and semantic errors were the most frequent errors on untrained items. Among the error types, semantic errors showed the largest change with sham treatment, and phonological errors showed the largest change with tDCS treatment. The statistical analysis showed that, averaging across the three time points, errors tended to decrease after tDCS for phonological nonword, phonological real-word, and unrelated errors. The strongest and statistically reliable effect was observed for phonological nonword errors at the two-month follow-up; immediate post-treatment and two-weeks post-treatment assessments showed numerically similar reductions that did not reach significance. For phonological real-word and unrelated errors, the pattern consistently favored tDCS over sham across time points (tDCS showed reductions, sham showed increases), but these contrasts did not meet the threshold for statistical significance at individual post-treatment time points. There was also a marginal increase in mixed errors after tDCS compared to a decrease after sham, driven by the large change at the two-month follow-up. The results suggest that cerebellar tDCS can reduce the frequency of certain error types on untrained items, with the most robust benefit emerging after a delay, consistent with consolidation-dependent plasticity (Jongkees et al., 2019).

The computerized aphasia treatment used in this study has been shown to improve naming in PWA in combination with tDCS (Baker et al., 2010; Fridriksson et al., 2011, 2018, 2019; Sebastian et al., 2020). By pairing pictures with spoken words and requiring rapid match/mismatch judgments across semantic foils, phonological foils, unrelated word foils, and targets, the task engages both semantic and phonological processing while imposing demands on lexical access, monitoring, and decision-making. Consistent with these demands, the largest improvements on untrained items were observed in phonological nonword and unrelated errors, which are most likely to benefit from repeated practice distinguishing target–word mappings from competing phonological and nonrelated alternatives. We also noted a marginal increase in mixed errors for untrained items. Because mixed errors are both phonologically and semantically related to the target, this pattern suggests increased interactivity among semantic and phonological nodes and indicates that the training exerts effects across multiple levels of the word-retrieval network. Although a reduction in semantic errors was anticipated given the task’s semantic requirements, we did not observe significant changes in semantic error rates on untrained items because semantic errors also declined in the sham condition, likely reflecting the impact of the active behavioral therapy, thus limiting power to detect a tDCS-specific effect on semantic error. In contrast, the clearest stimulation related benefit emerged for phonological nonword errors, consistent with a greater susceptibility of phonological encoding and monitoring processes to cerebellar modulation and with delayed gains that align with consolidation-dependent mechanisms (Reis et al., 2009).

The stimulation site, the right cerebellum, via cerebro-cerebellar loops, contributes to predictive timing, coordination, and optimization of lexical–semantic processing (Ackermann et al., 1998; Argyropoulos, 2016; De Smet et al., 2013; Murdoch, 2010; Strick et al., 2009). During therapy, participants repeatedly engaged in prediction, error monitoring, and rapid lexical search when deciding whether a spoken word matched a picture. Cerebellar tDCS may have augmented these processes, and in combination with training led to fewer phonological nonword and unrelated errors on untrained items, with benefits becoming most apparent after consolidation.

Dell’s Interactive Activation model posits that word retrieval involves interactive stages: lemma selection followed by phonological encoding with bidirectional feedback among semantic, lexical, and phonological nodes (Dell et al., 1997). Errors that are semantically or phonologically related to the target reflect a degree of residual integrity in the network, but rapid activation decay and competitive interference can lead neighbors to be selected in lieu of the intended item. From this perspective, the observed reductions in phonological nonword and unrelated errors after cerebellar tDCS plus computerized therapy suggest improved maintenance and stabilization of activation within the retrieval network, better control of competitive dynamics, and reduced noise. The delayed strengthening at two months for phonological errors is consistent with longer-term consolidation of network changes. By contrast, the absence of significant change in semantic errors aligns with prior reports showing limited impact on related error types following phonomotor intervention (e.g., Minkina et al., 2016, 2019). In Dell’s framework, intensive training that boosts phonological representations and their feedback can influence the broader network, but improvements may manifest first as reduced selection of clearly incorrect or nonword responses, and only later or with more targeted semantic training, as changes in semantically related errors. The consistent, albeit non-significant, tendency for phonological real-word errors to decrease after tDCS is compatible with the idea that enhanced feedback and sustained activation can strengthen competition resolution at the lemma-to-phonology interface, but the magnitude of change in our sample may have been insufficient to yield reliable timepoint-specific effects.

Polarity-specific analyses revealed no differential treatment effects between anodal and cathodal groups for any error type on the trained task, either when averaging across post-treatment timepoints or within individual timepoints. In contrast, for the untrained task (PNT), there was a significant Polarity Group (anode vs. cathode) × Treatment (tDCS vs. sham) interaction for phonological nonword errors when averaging across post-treatment timepoints. Within the anode group, phonological nonword errors decreased after tDCS and increased after sham, yielding a significant treatment effect. Planned timepoint comparisons indicated that this effect was driven primarily by a significant reduction at 2 months post-treatment, with marginal reductions at the immediate and 2-week post-treatment assessments. Within the cathode group, changes after tDCS and sham were not significant, and the overall treatment effect was not reliable. No other untrained error types showed polarity-specific treatment differences. Notably, these error-based findings differ from Sebastian et al., 2020 accuracy results, which suggested an advantage for cathodal cerebellar tDCS over anodal cerebellar tDCS (relative to sham). In the current study, the only reliable polarity effect favored anodal stimulation and was specific to reducing untrained phonological nonword errors, underscoring that these findings are consistent with the as-yet unclear directionality of the effects of anodal versus cathodal cerebellar tDCS on cognitive task performance (Grimaldi et al., 2016).

Taken together, cerebellar tDCS paired with computerized aphasia treatment did not differentially affect the error types for trained items relative to sham, but it reduced several untrained error types, with a robust, durable benefit for phonological nonword errors. This pattern suggests that combining noninvasive cerebellar stimulation with therapy may facilitate the broader language network and strengthen retrieval processes, yielding fewer errors after tDCS. This decrease in the various types of errors after tDCS may be due to the high duration and intensity of stimulation, which was 15 sessions, 3-5 times per week of tDCS (Sebastian et al., 2020). Repeated tDCS sessions administered simultaneously with behavioral therapy are believed to operate via mechanisms similar to long-term effectiveness, which is essential for neuroplasticity and memory consolidation (Fritsch et al., 2010; Reis et al., 2009, 2015).

Clinically, these results underscore the value of monitoring detailed error profiles rather than accuracy alone when implementing neuromodulatory adjuncts. Naming error analysis may bring greater insights about treatment responsiveness and mechanisms of change than naming accuracy data alone. These findings also highlight the importance of clinical error analysis for Speech-Language Pathologists in understanding the types and patterns of naming errors in PWA, which may help guide the pattern of recovery during the course of aphasia treatment.

## Limitations

These conclusions are tempered by modest, exploratory polarity subgroup samples and primary effects confined to a subset of error types, with mixed-error changes marginal and in need of replication. We focused on error-type changes to probe mechanism. Future studies should explicitly link error shifts to overall accuracy and functional communication outcomes to clarify clinical impact. Finally, treatment dosage and follow-up duration may have been insufficient to detect semantic-level changes, and larger, adequately powered trials are needed to confirm polarity effects and generalize findings across aphasia profiles. We are currently further analyzing more in-depth error coding analysis of the data for both trained and untrained naming in a larger randomized control trial (NCT05093673).

## Data Availability Statement

The data that support the findings of this study are available from the corresponding author, [RS], upon reasonable request.

## Supporting information

Supplemental Materials (Naming Errors)

## Data Availability

All data produced in the present study are available upon reasonable request to the authors

