## Supplemental Materials (Naming Errors) for "Investigating naming error patterns after non-invasive brain stimulation and language treatment in persons with aphasia"

**Figure S1. Participant flow diagram; Tx: Treatment; Ax: Assessment; 2 W: 2 weeks; 2 M: 2 months.**

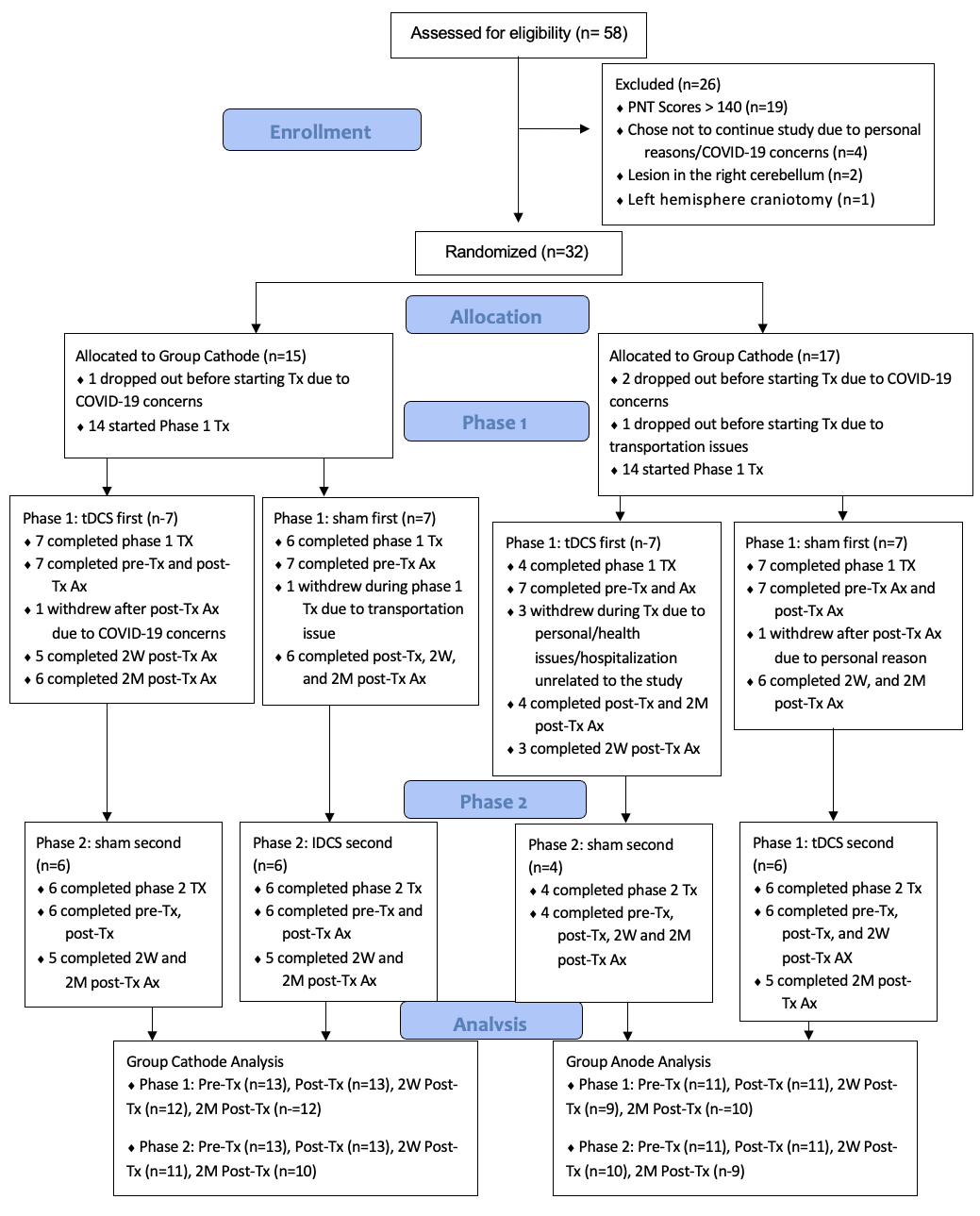

**Table S1a. Trained (N80) task. Post hoc tests for the Treatment x Timepoint interaction (timepoints changes relative to baseline).**

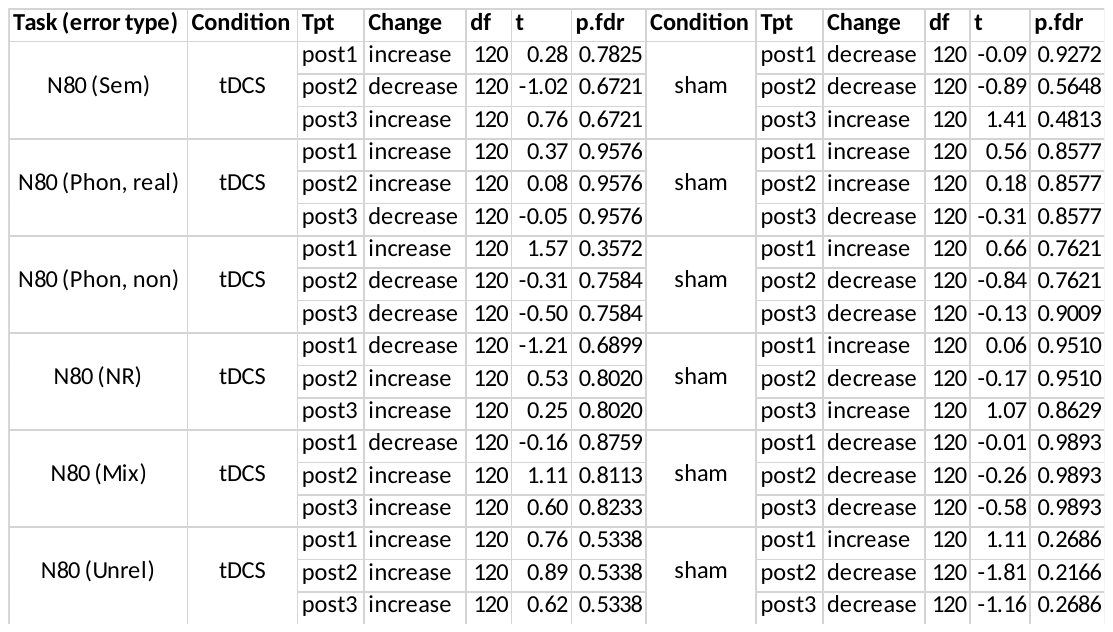

**Table S1b. Trained (N80) task. Post hoc tests for the Treatment x Timepoint interaction (tDCS vs sham, by timepoint).**

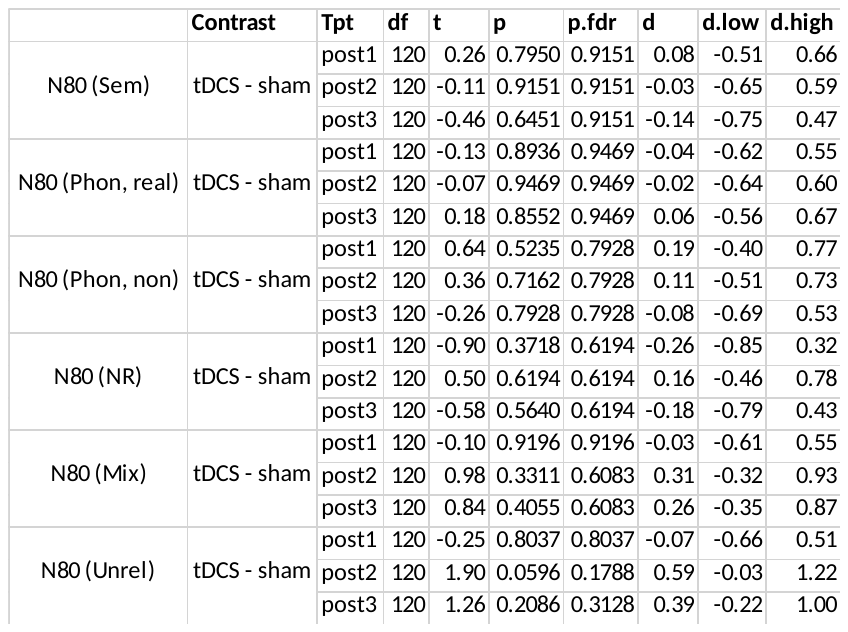

**Table S2a. Untrained (PNT) task. Post hoc tests for the Treatment x Timepoint interaction (timepoints changes relative to baseline).**

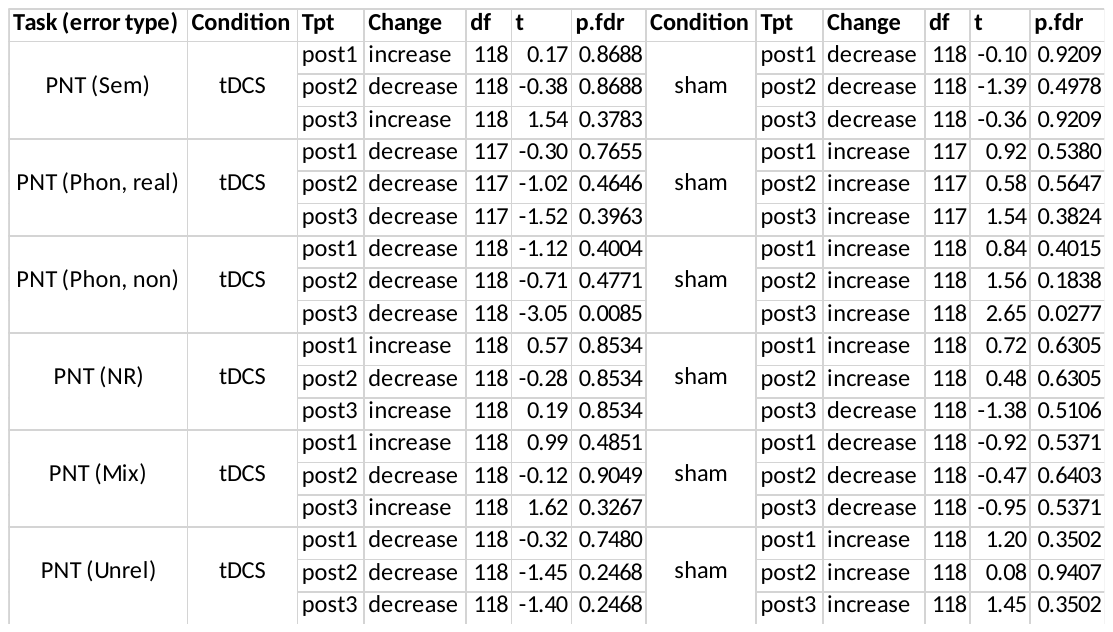

**Table S2b. Untrained (PNT) task. Post hoc tests for the Treatment x Timepoint interaction (tDCS vs sham, by timepoint).**

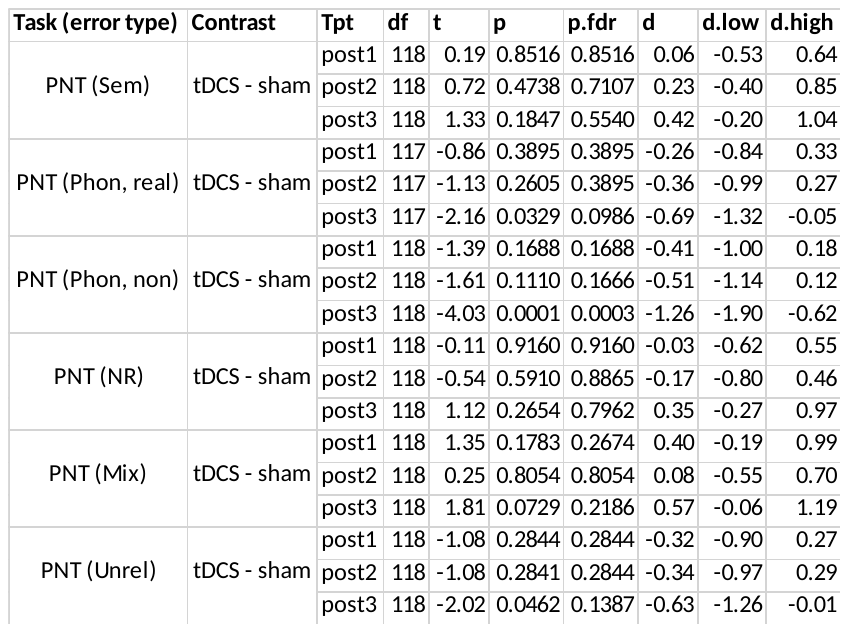

**Table S3a. Trained (N80) task. tDCS polarity group (Anodal). Post hoc tests for treatment effect (tDCS vs sham) for all timepoints.**

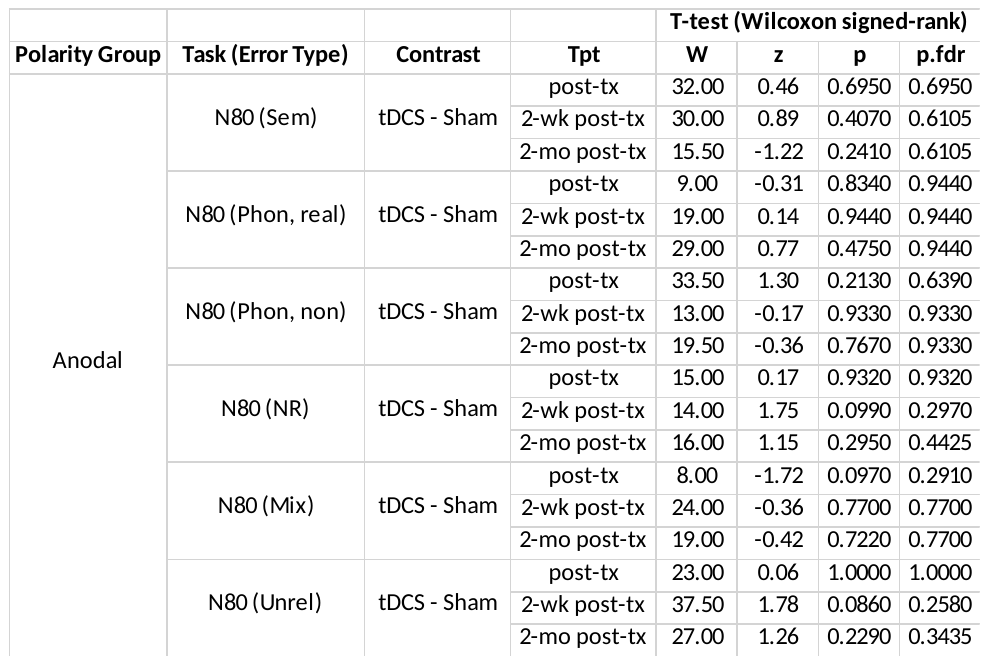

**Table S3b. Trained (N80) task. tDCS polarity group (Cathodal). Post hoc tests for treatment effect (tDCS vs sham) for all timepoints.**

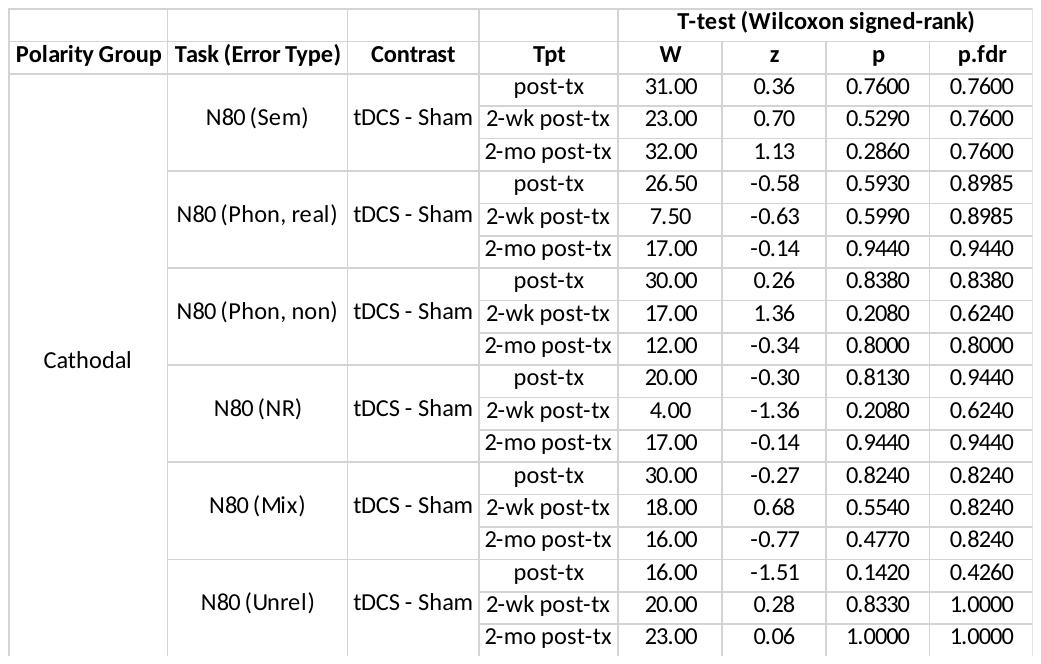

**Table S4a. Untrained (PNT) task. tDCS polarity group (Anodal). Post hoc tests for treatment effect (tDCS vs sham) for all timepoints.**

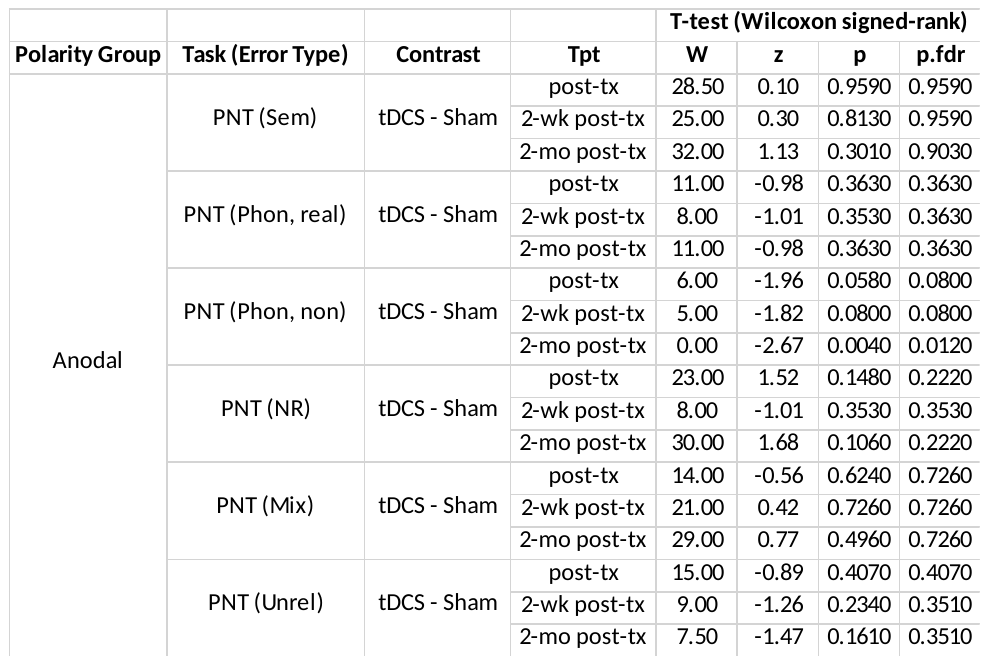

**Table S4b. Untrained (PNT) task. tDCS polarity group (Cathodal). Post hoc tests for treatment effect (tDCS vs sham) for all timepoints.**

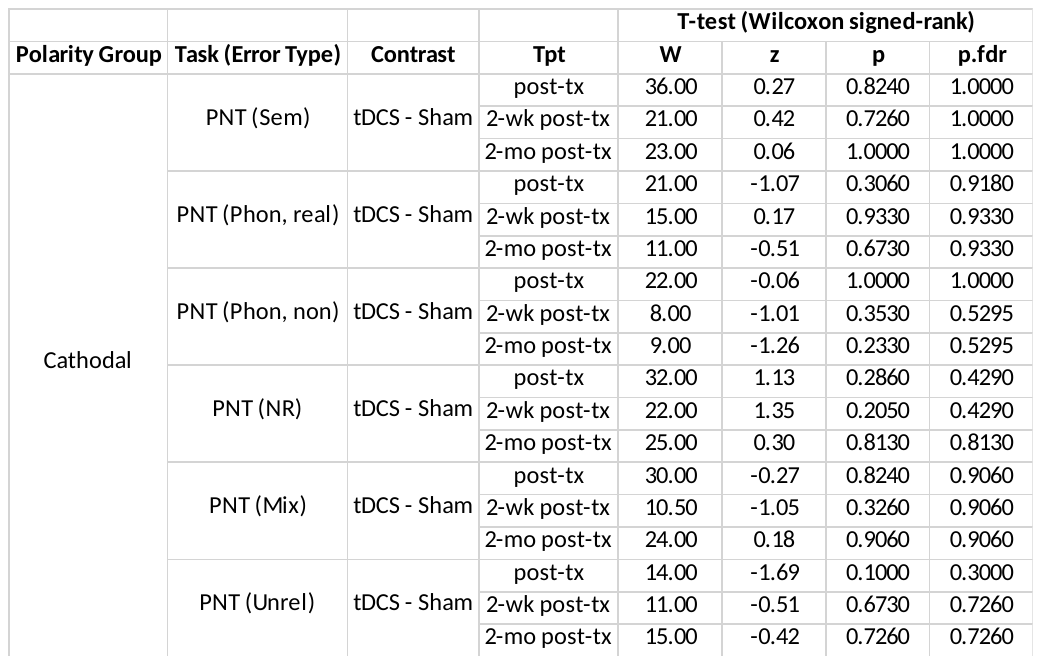
